# Mixed Chinese herbs and Western medicine for novel coronavirus disease 2019 (COVID-19): a mixed method review

**DOI:** 10.1101/2020.05.11.20098111

**Authors:** Zhang Yong Tai, Robby Miguel W.J Goh, Louisa Tay, Ruishu Zhou, Lin Ho Wong, Pang Ong Wong

## Abstract

**Background:** Coronavirus disease 2019 (COVID-19) is a pandemic affecting millions around the world. There is no existing pharmaceutical treatment that is known to be effective. Preliminary data shows that San Yao San Fang (SYSF) has clinical benefits in patients with COVID-19. The aim of this paper is to review existing data regarding the use of formulas within San Yao San Fang in the treatment of COVID-19

**Search Strategy:** We searched through EMBASE, Pubmed, Cochrane, Wanfang and China National Knowledge Infrastructure (CNKI) for studies on SYSF and patients with COVID-19 through April 2020.

**Eligibility Criteria:** We included studies that included a formula within San Yao San Fang with or without Western interventions against Western interventions.

**Main results:** We included 7 studies involving 532 patients. 3 retrospective observational studies and 1 randomised control trial reported on Lian Hua Qing Wen Jiao Nang, 1 randomised control trial on Jin Hua Qing Gan Ke Li, 1 retrospective observational study on Xue Bi Jing Zhu Se Ye and 1 randomised control trial on Qing Fei Pai Du Tang.

SYSF combined with Western interventions improved the recovery rate of symptoms such as fever (Risk Ratio (RR) 0.40 (95% CI 0.24 to 0.66, P < 0.01)), cough (RR 0.56 (95% CI 0.38 to 0.82, P < 0.01)) and fatigue (RR 0.61 (95% CI 0.47 to 0.78, P < 0.01)) and other symptoms such as headache, gastrointestinal symptoms, myalgia, dyspnoea and chest tightness (RR 0.63 (95% CI 0.47 to 0.83, P < 0.01)) as compared to the control group.

SYSF combined with Western interventions reduced the duration of fever as compared to the control group. (Mean difference (MD) -1.18 (95% CI -1.45 to -0.91, P < 0.01))

In regards to adverse events, there is no statistical difference between the treatment group treated with SYSF and Western interventions and the control group. (RR 1.62 (95% CI 0.83 to 3.17, P = 0.16)).

SYSF combined with Western interventions did not show to significantly reduce duration of hospitalisation as compared to the control group. (MD -0.73 (95% CI -5.19 to 3.73, P = 0.75))

**Conclusion:** SYSF appears to be clinically effective and safe. Further research is required to ensure the efficacy of SYSF.

## Introduction

Coronavirus disease 2019 (COVID-19) is a novel disease first identified in Wuhan, China, before spreading to the rest of the world.^1^ Since then, WHO has declared it as a Public Health Emergency of International Concern on the 30^th^ Jan 2020. COVID-19 patients are usually asymptomatic but could clinically manifest with flu-like symptoms like fever, fatigue, dry cough, anorexia, myalgia and dyspnea. In severe cases, it can progress to Acute Respiratory Distress Syndrome (ARDS) and death.^2,3^ Emerging studies have also demonstrated hypercoagulable states and Kawasaki disease as possible complications of COVID-19.^4,5^

Researchers in Wuhan have found death rates to be around 1%.^6^ The risk increases in patients who are older, male or who have other comorbidities like hypertension, diabetes, cardiovascular diseases and chronic lung diseases.^7^ As of 04 May 2020, the total number of confirmed deaths stands at 239,604.^8^ Till date, there is no definite treatment that has proven to be effective in the treatment of COVID-19. Remdesivir developed by GILEAD is one of the more promising medications for the treatment of COVID-19. A recent study done by Chinese researchers showed that patients did not benefit clinically from Remdesivir.^9^ It was however, refuted to be an underpowered trial which resulted in inconclusive findings.^10^

With the exponential rise in patients around the world, healthcare providers are under immense pressure to find a possible treatment to speed up the recovery of the patients and prevent a possible collapse of the healthcare system.

During the Severe Acute Respiratory Syndrome (SARS) epidemic in 2003, Traditional Chinese Medicine (TCM) practitioners found herbal medicine to be successful in the prevention and treatment of SARS.^11^ With this precedence, TCM practitioners are turning towards TCM in hopes of finding a treatment plan that can help prevent and treat COVID-19. TCM has been utilized with success in several reports for the treatment of COVID-19 pneumonia.^12^ More recently, a new class of herbal formulations known as San Yao San Fang (SYSF) (三药三方)) has been gaining widespread use for treatment in China.

SYSF refers to 3 existing formulas (San Yao) that are thought to be effective in the management of COVID-19 and 3 new formulations (San Fang) created during this pandemic for the treatment of COVID-19. San Yao includes 1. Jin Hua Qing Gan Ke Li (Formula J) (金花清感颗粒), ^13^ 2. Lian Hua Qing Wen Jiao Nang (Formula L) (连花清瘟胶囊)^14-15^ and 3. Xue Bi Jing Zhu Se Ye (Formula Xb) (血必净注射液).^16,17^

### How the intervention might work

Formula J and Formula L are oral herbal formulations that are typically used for flu like symptoms.^18^ Formula J targets inflammatory and apoptosis regulation by targeting various mechanisms such as PI3k-Akt, HIF-1, TNF, MAPK and NF-κB.^19^ On the other hand, Formula L works by inhibiting COVID-19 replication in Vero E6 cells and reduce production of pro-inflammatory cytokines such as TNF-α, IL-6, CCL-2/MCP-1 and CXCL-10/IP-10. ^20,21^

Formula Xb is an intravenous formulation typically used for fever, dyspnoea, palpitations and in cases like Systemic Inflammatory Response Syndrome (SIRS) and multi-organ failure.^22^ It has been employed in moderate-severe cases of COVID-19 in China.

San Fang includes 1. Hua Shi Bai Du Fang (Formula H) (化湿败毒方), 2. Qing Fei Pai Du Tang(Formula Q) (清肺排毒汤)^23-25^ and 3. Xuan Fei Bai Du Fang(Formula Xf) (宣肺败毒方).^26^ All 3 formulas were recently developed during the COVID-19 crisis to target the different clinical manifestations of COVID-19. Clinical trials are ongoing to identify the underlying mechanism behind the 3 different formulas.

### Objective of study

As early studies of SYSF seem to be promising, uncertainty remains about its effectiveness due to empirical usage in treatment of COVID-19 pneumonia. Thus, this study aims to review existing data regarding the use of formulas within SYSF in the treatment of COVID-19, and provide evidence for widespread recommendation and usage of SYSF as part of the management of COVID-19.

## Methods

### Criteria for considering studies for this review

#### Types of studies

Trials including the use of any of six Chinese formulas (1. Hua shi bai du fang 2. Jin Hua Qing Gan Ke Li 3. Lian hua qing wen jiao nang 4. Qing Fei Pai Du Tang 5. Xuan Fei Bai Du Tang 6. Xue Bi Jing Zhu She Ye) administered with or without Western medicines for COVID-19 patients were included.

#### Types of participants

COVID-19 cases diagnosed using any national diagnostic criteria, or by clinical impression of the physician-in-charge were included.

#### Types of interventions

We included trials administering

1. Hua shi bai du fang
2. Jin hua qing gan ke li
3. Lian hua qing wen jiao nang
4. Qing fei pai du tang
5. Xuan fei bai du tang
6. Xue bi jing zhe she ye

with or without Western medicines for COVID-19 patients, regardless of comparator.

#### Types of outcome measures

We assessed primary and secondary outcome measures at the end of treatment and/or at the end of follow-up.

#### Primary outcomes

1. Persistence of fever, cough and fatigue post0 treatment

#### Secondary outcomes

1. Persistence of other symptoms (excluding fever, cough and fatigue) post treatment
2. Duration of each symptom.
3. Number of days in hospital
4. Adverse events/side effects
5. Hospitalization rate

### Search methods for identification of studies

#### Electronic searches

For this review we searched EMBASE, Pubmed, Cochrane, Wanfang and China National Knowledge Infrastructure (CNKI) (2019-2020)

### EMBASE, Pubmed And Cochrane

1. Jin hua qing gan AND Covid OR Coronavirus
2. Lian hua qing wen AND Covid OR Coronavirus
3. Xue bi jing zhu AND Covid OR Coronavirus
4. Qing fei pai du tang AND Covid OR Coronavirus
5. Xuan fei bai du tang AND Covid OR Coronavirus
6. Xue bi jing zhu AND Covid OR Coronavirus
7. Traditional Chinese Medicine AND Covid OR Coronavirus

### CNKI (overseas CNKI) And Wanfang (based on the translations of the keywords used in English medium)

1. 金花清感颗粒 AND 新冠状病毒/冠状病毒/新冠肺炎
2. 连花清瘟胶囊 AND 新冠状病毒/冠状病毒/新冠肺炎
3. 血必净注射液 AND 新冠状病毒/冠状病毒/新冠肺炎
4. 化湿败毒方 AND 新冠状病毒/冠状病毒/新冠肺炎
5. 清肺排毒汤 AND 新冠状病毒/冠状病毒/新冠肺炎
6. 宣肺败毒方 AND 新冠状病毒/冠状病毒/新冠肺炎
7. 中药 AND 新冠状病毒/冠状病毒/新冠肺炎

#### Searching other resources

We did not impose any language or publication restrictions.

### Data collection and analysis

#### Selection of studies

Four review authors (ZY, L, RS, RM) independently reviewed the titles, abstracts and keywords of all records retrieved to determine the studies to be assessed. We retrieved full articles for further assessment if the information given suggested that the study:

1. Included COVID-19 patients;
2. And administered any of six Chinese herbs (1. Hua shi bai du fang 2. Jin hua qing gan ke li 3. Lian hua qing wen jiao nang 4. Qing fei pai du tang 5. Xuan fei bai du tang 6. Xue bi jing zhu she ye) for the treatment of COVID-19

A fifth review author (LH) acted as arbiter and resolved any differences in opinion.

#### Data extraction and management

Four review authors (ZY, L, RS, RM) independently extracted data from each included trial using a standard extraction form, which included the following items.

1. General information: published/unpublished, language, authors, article title, journal title and year, volume, issue, page, funding source.
2. Design of the trial: prescribed size, generation of randomisation sequence, allocation concealment method, blinding information, statistical methods and attrition.
3. Participants: diagnostic criteria, total number and number in comparison groups, baseline characteristics, age, gender, inclusion criteria, exclusion criteria, study setting.
4. Intervention: type of herbs and (if any) Western medicine, the content of herbal formulas, duration, times, dose, co-intervention, control, withdrawals, drop out, loss to follow-up.
5. Outcome: all outcomes.
6. Conclusion: positive/negative.

### Assessment of risk of bias in included studies

We assessed risk of bias following the recommendations in the *Cochrane Handbook for Systematic Reviews of Interventions*.^27^ We used a #x2018;Risk of bias’ table to assess the methodological quality of the trials.

1. Sequence generation: describes the method used to generate the allocation sequence in sufficient detail to allow an assessment of whether it should produce
2. Allocation concealment: describes the method used to conceal the allocation sequence in sufficient detail to determine whether intervention allocations could have been foreseen in advance of, or during, enrolment.
3. Blinding of participants, personnel and outcome assessors. Assessments should be made for each main outcome (or class of outcomes): describes all measures used, if any, to blind study participants and personnel from knowledge of which intervention a participant received. Provide any information relating to whether the intended blinding was effective.
4. Incomplete outcome data. Assessments should be made for each main outcome (or class of outcomes): describes the completeness of outcome data for each main outcome, including attrition and exclusions from the analysis. State whether attrition and exclusions were reported, the numbers in each intervention group (compared with total randomised participants), reasons for attrition/exclusions where reported, and any reinclusions in analyses performed by the review authors.
5. Selective outcome reporting: state how the possibility of selective outcome reporting was examined by the review authors, and what was found.
6. Other sources of bias: state any important concerns about bias not addressed in the other domains in the tool. If particular questions/entries were pre-specified in the review’s protocol, responses should be provided for each question/entry.

Two review authors (ZY, LH) independently assessed each trial. We resolved disagreements by discussion.

### Measures of treatment effect

We extracted both dichotomous data and continuous data with 95% confidence intervals (CIs). We used risk ratios (RR) for dichotomous data. We calculated mean differences (MD) for continuous data. We calculated overall results based on the random-effects model if heterogeneity existed between studies. If no heterogeneity was detected between studies, we used the fixed-effect model.

### Unit of analysis issues

We analysed the data using Review Manager (RevMan 2011) software. We summarised data statistically if they were available and of sufficient quality and similarity. We performed meta-analyses within comparisons where individual trials compared the same trial intervention versus the same control intervention.

## Results

### Description of studies

#### Results of the search

The initial search of electronic databases yielded 476 studies. After scanning the results, 3 RCT and 4 observational trials were identified, which appeared to meet the inclusion criteria as shown in Figure 1.

**Figure 1.**
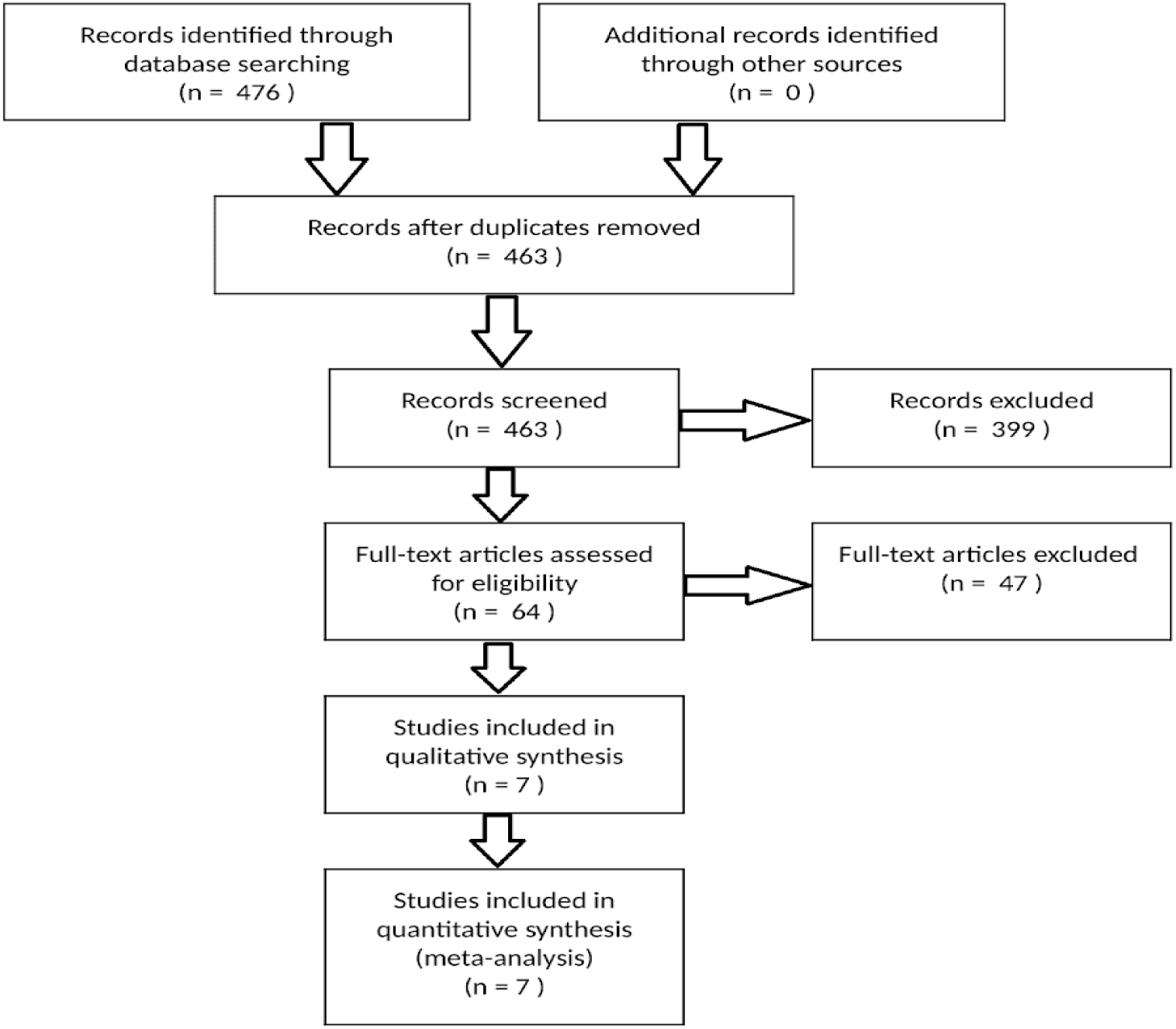
Study flow diagram

#### Assessment of heterogeneity

We tested heterogeneity using the Z score and the Chi^2^ test with significance being set at P < 0.1 and planned to explore possible sources of heterogeneity by subgroup and sensitivity analysis.

#### Data synthesis

We calculated overall results based on the random-effects model if heterogeneity existed between trials. If no heterogeneity was detected between studies, we considered the fixed-effect model. Hypothesis tests used the Z test. We considered the results had achieved statistical significance if P ≤ 0.1. If P > 0.1, we considered that the results had not achieved statistical significance. CIs were set at 95%.

#### Subgroup analysis and investigation of heterogeneity

We planned to do a subgroup analysis between RCTs and retrospective observational trials.

#### Included trials

Seven studies were included for meta-analysis - Cheng et al., 2020 ^28^; Duan et al., 2020 ^13^; Lv et al., 2020 ^29^; Li et al., 2020 ^24^; Wang et al., 2020 ^30^;Yao et al., 2020 ^31^; and Zheng et al., 2020 ^16^. Contents of the formulas and their commonly used indications are specified in Table 1. Summary of the trials are shown in Tables 2-5.

**Table 1.** SYSF formulas and respective herb constituents

| Formula<br>English<br>name | Herb constituents Scientific name | Herb constituents<br>Chinese name | Indications |
| --- | --- | --- | --- |

| Chinese name |  |  |  |
| --- | --- | --- | --- |
| Hua Shi Bai Du Fang<br>化湿败毒方<br>(Formula H) | Ephedra sinica Stapf, Herba Pogostemonis, Gypsum Fibrosum, Semen Armeniacae, Pinellia ternata (Thunb.) Breit, Cortex Magnoliae Officinalis, Rhizoma Atractylodis, Fructus Tsaoako, Poria, Radix Astragali, Radix Paeoniae Rubra, Semen Lepidii, Radix et Rhizoma Rhei, Radix Glycyrrhizae | 生麻黄、藿香、生石膏、杏仁、法半夏、厚朴、苍术、草果、茯苓、生黄芪、赤芍、葶苈子、生大黄、甘草 | Mild-moderate COVID-19 |
| Jin Hua Qing Gan Ke Li<br>金花清感颗粒<br>(Formula J) | Flos Lonicerae, Gypsum Fibrosum, Ephedra sinica Stapf, Semen Armeniacae, Radix Scutellariae, Fructus Forsythiae, Bulbus Fritillariae Thunbergii, Rhizoma Anemarrhenae, Arctium lappa L., Herba Artemisiae Annuae, Herba Menthae Haplocalycis, Radix Glycyrrhizae | 金银花、石膏、麻黄、苦杏仁、黄芩、连翘、浙贝母、知母、牛蒡子、青蒿、薄荷、甘草 | Commonly used:<br>Flu like symptoms<br>Mild-moderate COVID-19 |
| Lian Hua Qing Wen Jiao Nang<br>连花清瘟胶囊<br>(Formula L) | Fructus Forsythiae, Flos Lonicerae, Ephedra sinica Stapf, Rhizoma Dryopteris, Radix Isatidis, Gypsum Fibrosum, Herba Menthae Haplocalycis, Herba Pogostemonis, Rhodiola rosea L., Herba Houttuyniae, Radix et Rhizoma Rhei, Semen Armeniacae, Radix Glycyrrhizae | 连翘、金银花、炙麻黄、绵马贯众、板蓝根、石膏、薄荷脑、广藿香、红景天、鱼腥草、大黄、炒苦杏仁、甘草 | Commonly used:<br>Flu like symptoms<br>Mild-moderate COVID-19 |
| Qing Fei Pai Du Tang<br>清肺排毒汤<br>(Formula Q) | Herba Ephedrae, Radix Glycyrrhizae Preparata, Semen Armeniacae, Gypsum Fibrosum, Ramulus Cinnamomi, Rhizoma Alismatis, Polyporus, Rhizoma Atractylodis Macrocephalae, Poria, Radix Bupleuri, Radix Scutellariae, Rhizoma Pinelliae Preparatum, Rhizoma Zingiberis Recens, Radix Asteris, Tussilago farfara, Rhizoma Belamcandae, Herba cum Radix Asari, Rhizoma Dioscoreae, Fructus Aurantii Immaturus, Pericarpium Citri Reticulatae, Herba Pogostemonis | 麻黄、炙甘草、杏仁、生石膏、桂枝、泽泻、猪苓、白术、茯苓、柴胡、黄芩、姜半夏、生姜、紫菀、冬花、射干、细辛、山药、枳实、陈皮、藿香 | Mild-moderate COVID |
| Xuan Fei<br>Bai Du<br>Fang<br>宣肺败毒<br>方<br>(Formula<br>Xf) | Herba Ephedrae, Semen Armeniacae,<br>Gypsum Fibrosum, Semen Coicis,<br>Rhizoma Atractylodis, Herba<br>Pogostemonis, Herba Artemisiae<br>Annuae, Reynoutria japonica Houtt.,<br>Herba Verbenae, Rhizoma Imperatae,<br>Semen Lepidii, Fructus Citri Grandis,<br>Radix Glycyrrhizae | 麻黄、苦杏仁、生石<br>膏、生薏苡仁、茅苍<br>术、广藿香、青蒿草、<br>虎杖、马鞭草、干茅<br>根、葶苈子、化橘红、<br>生甘草 | Mild-moderate<br>COVID-19 |
| Xue Bi<br>Jing Zhu<br>She Ye<br>血必净注<br>射液<br>(Formula<br>Xb) | Flos Carthami, Radix Paeoniae Rubra,<br>Rhizoma Chuanxiong, Radix Salviae<br>Miltiorrhizae, Radix Angelicae Sinensis | 红花、赤芍、川芎、<br>丹参、当归 | Commonly used:<br>Fever, dyspnoea,<br>palpitations,<br>presences of<br>systemic<br>inflammatory<br>response and multi-organ failure.<br>Moderate-severe<br>COVID-19 |

**Table 2.** Summary of existing evidence of Jin Hua Qing Gan Ke Li in the treatment of Covid-19

|  |  |
| --- | --- |
| Author, year | Duan, 2020 |
| Type of study | RCT |
| Sample size | 123<br>Treatment group (n=82) and Control group (n=41) |
| Intervention<br>duration (days) | 5 |
| Treatment<br>group<br>intervention | Jinhua Qinggan granules administered at dosage of 5g/ bag, 2 bags each time, 3<br>times a day |
| Control group<br>intervention | Conventional western treatment including symptomatic treatment such as anti-viral<br>and anti-infection |
| Outcomes | Recovery rate of fever, fatigue, cough and sputum symptoms in the treatment group were significantly higher than those in the control group.<br>There were 27 cases (32.93%) of adverse reactions in the treatment group, all of which were diarrhoea, 8 patients discontinued the drug due to intolerance, symptoms of diarrhoea improved after the withdrawal.<br>14 cases fell out of the Treatment group (8 cases due to gastrointestinal adverse reactions, 3 cases due to drug taste problems, 3 cases of uncooperative treatment). 68 cases of final effective cases, of which 9 patients were admitted to the hospital due to aggravated illness but continued Jinhua Qinggan granules intake. 19 patients reported diarrhoea after taking the medicine or worsened diarrhoea. Total of 7 cases were lost in the Control group (due to uncooperative treatment), 34 cases were effective, of which 10 cases were admitted to the hospital due to aggravated illness. No adverse reaction in the control group. This difference between the treatment and control group is statistically significant |

**Table 3.** Summary of existing evidence of Lian Hua Qing Wen Jiao Nang in the treatment of Covid-19

| Author, year | Cheng et al, 2020 | Wang et al, 2020 | Lv et al, 2020 | Yao et al, 2020 |
| --- | --- | --- | --- | --- |
| Type of study | Retrospective Observational Study | RCT | Retrospective Observational Study | Retrospective Observational Study |
| Sample size | 102<br>Treatment Group (n=51) and Control Group (n=51) | 60<br>Treatment group (n=30) and Control Group (n=30) | 101<br>Treatment group (n=63) and Control group (n=38) | 42<br>Treatment group (n=21) and Control group (n=21) |
| Intervention duration (days) | 7 | 7-10 | 10 | 20 |
| Treatment group intervention | Conventional western treatment with Lianhua Qingwen granules, 6g/bag, 3 times/ day | Lianhua Qingwen capsules 4 pieces, 3 times/ day, interferon-α 500u nebulised, 2 times/ day | Lianhua Qingwen granules, 6g/ packet, 3 times/ day. IV Moxifloxiacin 400mg once/ day, IV Ganciclovir 500mg IV once/ day, IVIg 2.5g once/ day, IV Ambroxol 30mg 2 times/ day, IV Doxofylline 200mg once/ day, IV Methylprednisolone 40mg once/ day | Basic treatment (interferon-α 500u nebulised, Lopinavir 200mg/ ritonavir 50mg 2 tablets each, Ribavirin IV 500mg and Chloroquine 500mg 2 times/ day. Umifenovir 200mg 3 times/ day) in combination with Lianhua Qingwen granules, 1 packet, 3 times/day |
| Control group intervention | Conventional western treatment including symptomatic treatment such as anti-virus and anti-infection | Lopinavir/ritonavir | IV Moxifloxiacin 400mg once/ day, IV Ganciclovir 500mg IV once/ day, IVIg 2.5g once/ day, IV Ambroxol 30mg 2 times/ day, IV Doxofylline 200mg once/ day, IV Methylprednisolone 40mg once/ day | Basic treatment (interferon-α 500u nebulised, Lopinavir 200mg/ ritonavir 50mg 2 tablets each, Ribavirin IV 500mg and Chloroquine 500mg 2 times/ day. Umifenovir 200mg 3 times/ day) |
| Outcomes | In terms of recovery rate of symptoms, the treatment group showed faster recovery that is statistically significant.<br>The control group has a greater incidence of worsening of symptoms that is statistically significant.<br>In terms of improvement in CT Thorax, there is no statistical significant difference between the 2 groups. | Outcome is based on the effectiveness of protocol on major criteria and minor criteria. Major criteria include resolution of fever and imaging tests, and normalisation of CRP and lymphocyte count. Minor Criteria include resolution of cough, fatigue, chest tightness and normalisation of neutrophil count and ESR. Major effectiveness includes meeting all major criteria. Minor effectiveness includes meeting at least 1 major criteria and any minor criteria. Treatment group achieved 76.67% all-effectiveness, as compared to the 46.67% all-effectiveness of the control group, making it statistically significant. | The Treatment group has a statistically significant recovery rate in fever, cough and fatigue as compared to the control group. There is no statistical difference in the recovery of other symptoms in both Treatment and Control groups.<br>There is no statistical significant difference in adverse events in both Treatment and Control groups. | Patients in the Treatment group had higher clinical effect as compared to the Control group, including recovery rate of cardinal symptoms like fever (85.7% vs 57.1%), cough (46.7% vs 5.6%), expectoration (64.3% vs 9.1%) and shortness of breath (77.8% vs 0%). Duration of fever shortened by 1.5 days in the Treatment group. |

**Table 4.** Summary of existing evidence of Qing Fei Pai Du Tang in the treatment of Covid-19

|  |  |
| --- | --- |
| Author, year | Li et al, 2020 |
| Type of study | RCT |
| Sample size | 60<br>Treatment group (n=30) and Control group (n=30) |
| Intervention duration (days) | 44 |
| Treatment group intervention | Antiviral treatment including Lopinavir/ritonavir, Umifenovir and Oseltamivir, and Modified Qingfei Paidu Decoction |
| Control group intervention | Antiviral treatment including Lopinavir/ritonavir, Umifenovir and Oseltamivir |
| Outcomes | Hospitalization time (13.6 vs 16.4 days), resolution of fever (2.3 vs 3.9 days) and cough (4.9 vs 6.5 days) of the Treatment group was significantly lower than in the Control group. Complete recovery rate of the Treatment group was 90% compared to 83.33% in the Control group.<br>There is no statistical difference in terms of worsening of patient condition between the Control and Treatment group.<br>Time taken for improvement of CT Thorax in Treatment group was also significantly shortened (6.6 vs 8.8 days). One case of a patient with an adverse reaction observed in the Treatment Group as compared to 3 in the Control group, which improved with pantoprazole. |

**Table 5.** Summary of existing evidence of Xue Bi Jing Zhu She Ye in the treatment of Covid-19

|  |  |
| --- | --- |
| Author, year | Zhang et al, 2020 |
| Type of study | Retrospective Observational Study |
| Sample size | 44<br>Treatment group (n=22) and Control group (n=22) |
| Intervention duration (days) | 4-25 |
| Treatment group intervention | Routine antiviral (interferon-α nebulised and oral Umifenovir granule) and symptomatic supportive treatment, with IV drip (50 mL Xuebijing injection dissolved in 100 mL saline solution), twice a day |
| Control group intervention | Routine antiviral (interferon-α nebulised and oral Umifenovir granule) and symptomatic supportive treatment |
| Outcomes | The effective rate of lung CT absorption (95.5%) and overall treatment (68.2%) in the Treatment group were higher than those in the Control group ($P < 0.05$), shows Xuebijing injection could reduce the occurrence of severe cases, and has guiding significance for clinical treatment. No significant difference in the recovery level of inflammatory indexes (WBC, L, L%, CRP, ferritin) and the negative conversion rate of nucleic acid between the two groups before and after treatment ($P > 0.05$). The incidence of adverse reactions in the Treatment group was 9.10%, compared with 4.55% of the Control group, the difference was not statistically significant. Neither group experienced serious adverse reactions, symptomatic treatment can alleviate the symptoms of diarrhea, nausea and dry cough observed. |

#### Excluded trials

13 records were duplicates and were removed after the initial search. 447 of the remaining studies were removed because they were either

1. Discussing the pharmacological properties of SYSF and not clinical studies,
2. Discussing how SYSF would theoretically work, or
3. Case reports on individual patients

### Risk of bias in included studies

#### Allocation

The three RCT trials (Duan et al., Cheng et al., Yao et al.) described the methods used to generate random allocation sequences. The other four trials did not mention how allocation was conducted.

#### Blinding

Single blinding was used in all three of the RCTs (Duan et al., Wang et al., Li et al.).

#### Selective reporting

Wang et al. reported an improvement by using a criteria which includes symptoms, clinical signs and laboratory results, there were no further breakdown of the data.

#### Other potential sources of bias

No other potential sources of bias were identified.

#### Effects of intervention

##### 1. Persistence of symptoms post treatment

###### a. Fever

Three studies reported this outcome and were analysed - Cheng et al., 2020; Lv et al., 2020 and Yao et al., 2020.

The total population analysed comprised 220 patients, 124 in the SYSF and Western medicine arm, and 96 in the Western medicine alone arm. This gave a statistically significant risk ratio of 0.40 (95% CI 0.24 to 0.66, P < 0.01) in favour of the SYSF and Western medicine arm, with an I2 of 0% (Fig. 2), suggesting that patients receiving SYSF and Western medicine recover from fever sooner after the initiation of treatment.

**Figure 2.**
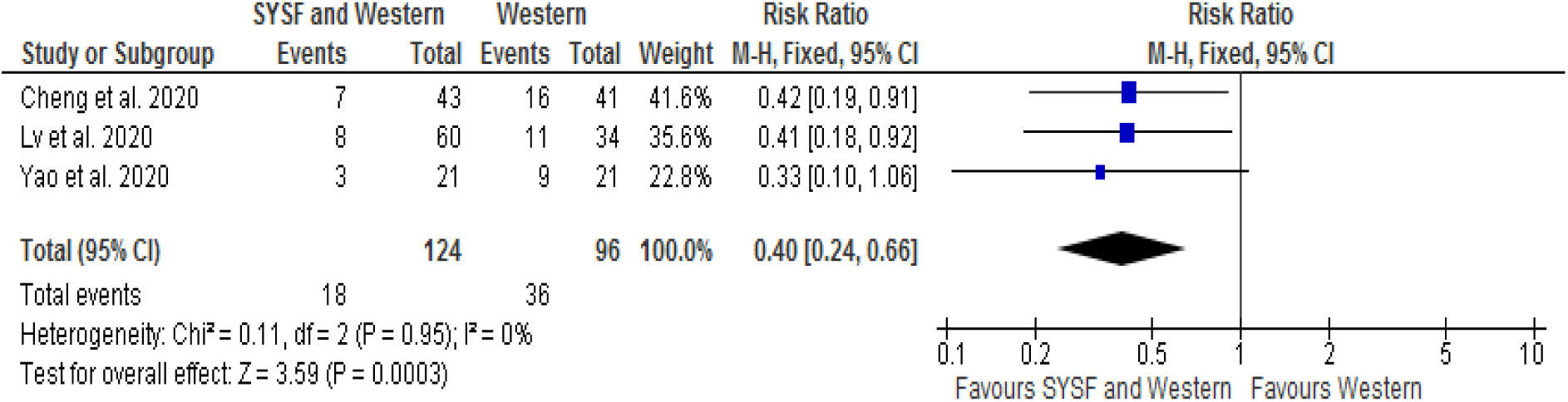
Persistence of fever post treatment between combined therapy vs Western treatment alone

###### b. Cough

Three studies reported this outcome and were analysed - Cheng et al., 2020; Lv et al., 2020 and Yao et al., 2020.

The total population analysed comprised 177 patients, 96 in the SYSF and Western medicine arm, and 81 in the Western medicine alone arm. This gave a statistically significant risk ratio of 0.61 (95% CI 0.47 to 0.78, P < 0.01)in favour of the SYSF and Western medicine arm, with an I2 of 0% (Fig. 3), suggesting that patients receiving SYSF and Western medicine recover from cough sooner after the initiation of treatment.

**Figure 3.**
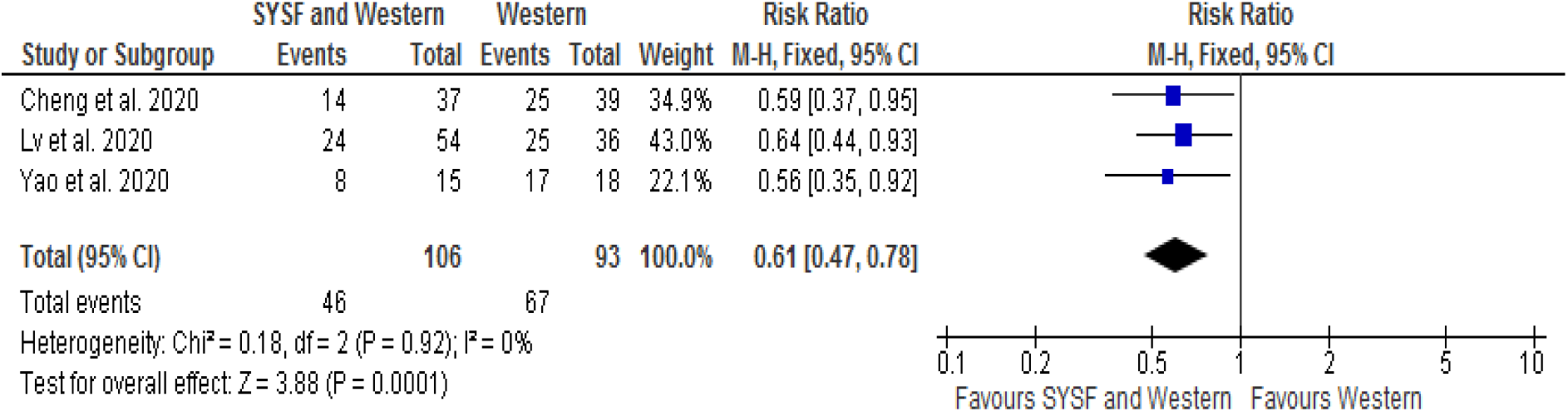
Persistence of cough by post treatment between combined therapy vs Western treatment alone

###### c. Fatigue

Three studies reported this outcome and were analysed - Cheng et al., 2020; Lv et al., 2020 and Yao et al., 2020.

The total population analysed comprised 199 patients, 106 in the SYSF and Western medicine arm, and 93 in the Western medicine alone arm. This gave a statistically significant risk ratio of 0.56 (95% CI 0.38 to 0.82, P < 0.01) in favour of the SYSF and Western medicine arm, with an I2 of 28% (fig. 4), suggesting that patients receiving SYSF and Western medicine recover from fatigue sooner after the initiation of treatment.

**Figure 4.**
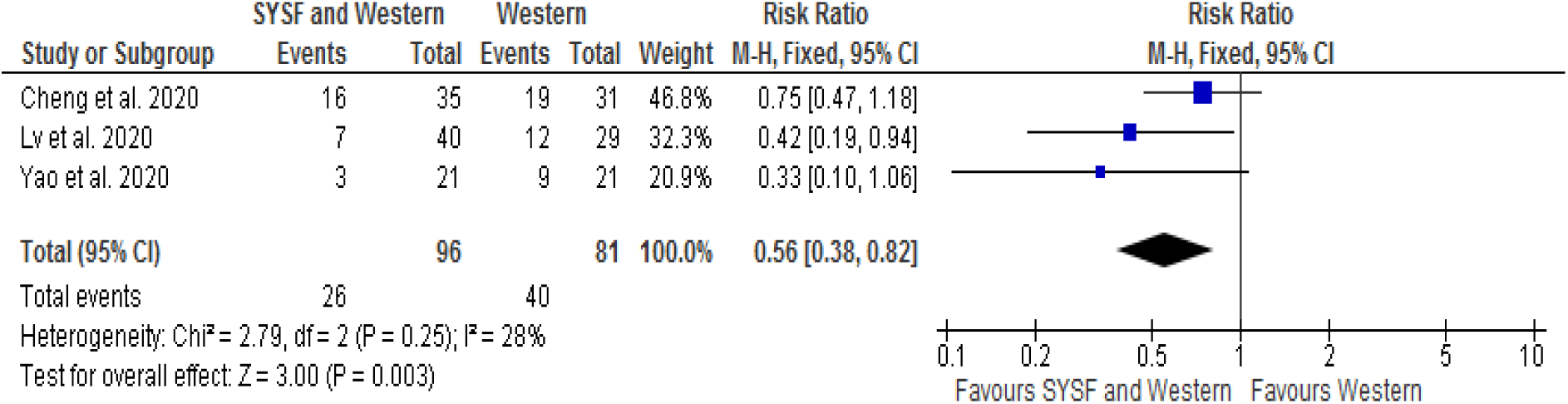
Persistence of fatigue post treatment between combined therapy vs Western treatment alone

##### 2. Duration of fever

Four studies reported this outcome and were analysed - Cheng et al., 2020; Li et al., 2020; Lv et al., 2020 and Zhang et al., 2020.

The total population analysed comprised 259 patients, 148 in the SYSF and Western medicine arm, and 111 in the Western medicine alone arm. This gave a statistically significant mean difference of -1.18 (95% CI -1.45 to -0.91, P < 0.01) in favour of the SYSF and Western medicine arm, with an I2 of 46% (fig. 5), suggesting that patients receiving SYSF and Western medicine had fevers for a shorter duration.

**Figure 5.**
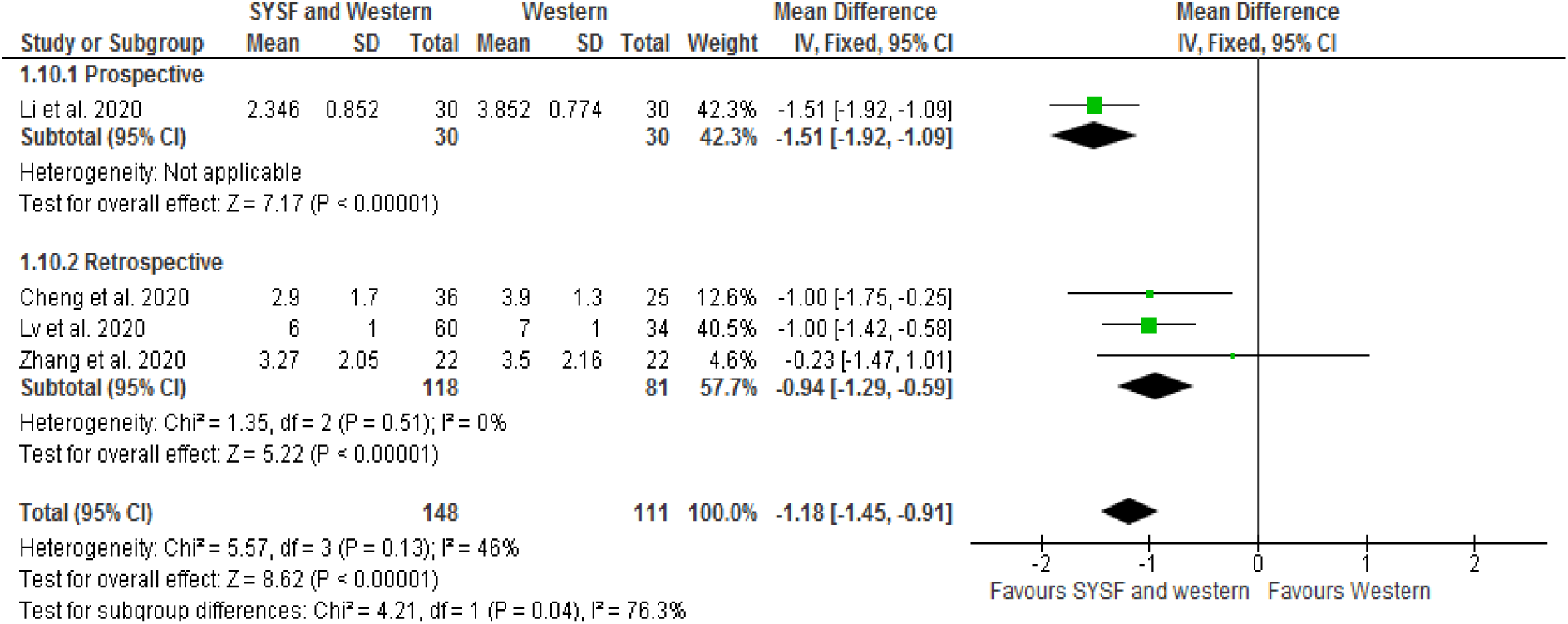
Duration of fever between combined therapy vs Western treatment alone.

In addition, we performed subgroup analyses of prospective and retrospective studies separately.

The prospective studies comprised only Li et al., 2020 thus meta-analysis was not conducted. This study had 60 patients, 30 in the SYSF and Western medicine arm, and 30 in the Western medicine alone arm. This gave a statistically significant mean difference of -1.51 (95% CI -1.92 to -1.09, P < 0.01) in favour of the SYSF and Western medicine arm (Fig. 5), suggesting that patients participating in studies performed prospectively and received SYSF and Western medicine had fevers for a shorter duration.

The retrospective studies analysed were Cheng et al., 2020; Lv et al., 2020 and Zhang et al., 2020 comprising 199 patients, 118 in the SYSF and Western medicine arm, and 81 in the Western medicine alone arm. This gave a statistically significant mean difference of -0.94 (95% CI -1.29 to -0.59, P < 0.01) in favour of the SYSF and Western medicine arm, with an I2 of 46% (Fig. 5), suggesting that patients who participated in studies performed retrospectively and received SYSF and Western medicine had fevers for a shorter duration.

##### 3. Persistence of other symptoms post treatment

Five studies reported this outcome and were analysed - Cheng et al., 2020; Duan et al., 2020; Lv et al., 2020; Wang et al., 2020 and Yao et al., 2020.

The total population analysed comprised 1279 patients, 762 in the SYSF and Western medicine arm, and 517 in the Western medicine alone arm. This gave a statistically significant risk ratio of 0.63 (95% CI 0.47 to 0.83, P < 0.01) in favour of the SYSF and Western medicine arm, with an I2 of 79% (Fig. 6), suggesting that patients receiving SYSF and Western medicine had symptoms such as headache, gastrointestinal symptoms, myalgia, dyspnea and chest tightness for a shorter duration.

**Figure 6.**
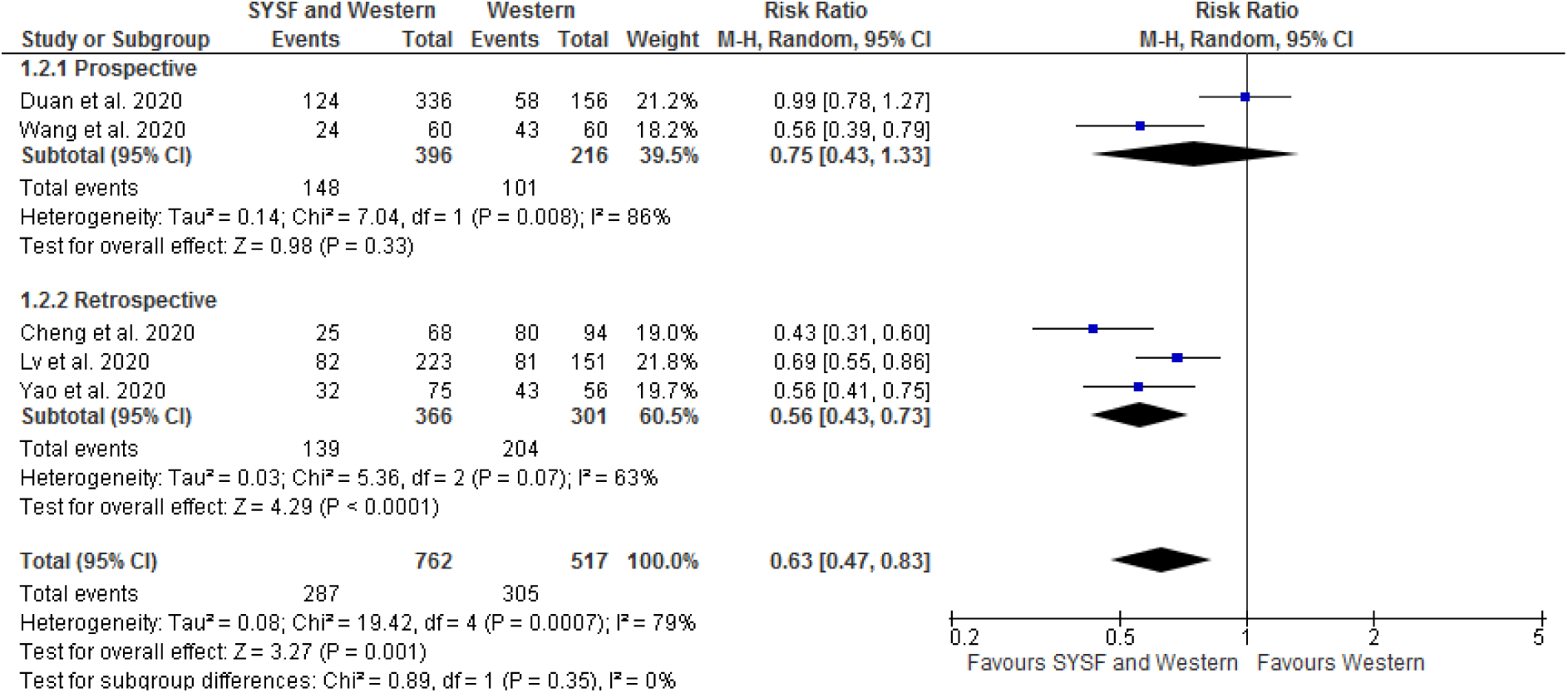
Persistence of other symptoms post treatment between combined therapy vs Western treatment alone.

In addition, we performed subgroup analyses of prospective and retrospective studies separately.

The prospective studies analysed were Duan et al., 2020 and Wang et al., 2020 comprising 612 patients, 396 in the SYSF and Western medicine arm, and 216 in the Western medicine alone arm. This gave a risk ratio of 0.75 (95% CI 0.43 to 1.33, P = 0.33) with an I2 of 86% (Fig. 6), suggesting no statistically significant difference in the persistence of symptoms between the two arms in studies performed prospectively.

The retrospective studies analysed were Cheng et al., 2020; Lv et al., 2020 and Yao et al., 2020 comprising 667 patients, 366 in the SYSF and Western medicine arm, and 301 in the Western medicine alone arm. This gave a statistically significant risk ratio of 0.56 (95% CI 0.43 to 0.73, P < 0.01) in favour of the SYSF and Western medicine arm, with an I2 of 63% (Fig. 6), suggesting that patients who participated in studies analysed retrospectively and received SYSF and Western medicine had symptoms such as headache, gastrointestinal symptoms, myalgia, dyspnea and chest tightness for a shorter duration.

##### 4. Adverse reactions

Four studies reported this outcome and were analysed - Duan et al., 2020; Li et al., 2020; Lv et al., 2020 and Zhang et al., 2020.

The total population analysed comprised 328 patients, 197 in the SYSF and Western medicine arm, and 131 in the Western medicine alone arm. This gave a risk ratio of 1.62 (95% CI 0.83 to 3.17, P = 0.16) with an I2 of 80% (Fig. 7), suggesting that there is no statistically significant difference in reported adverse reactions between the two arms.

**Figure 7.**
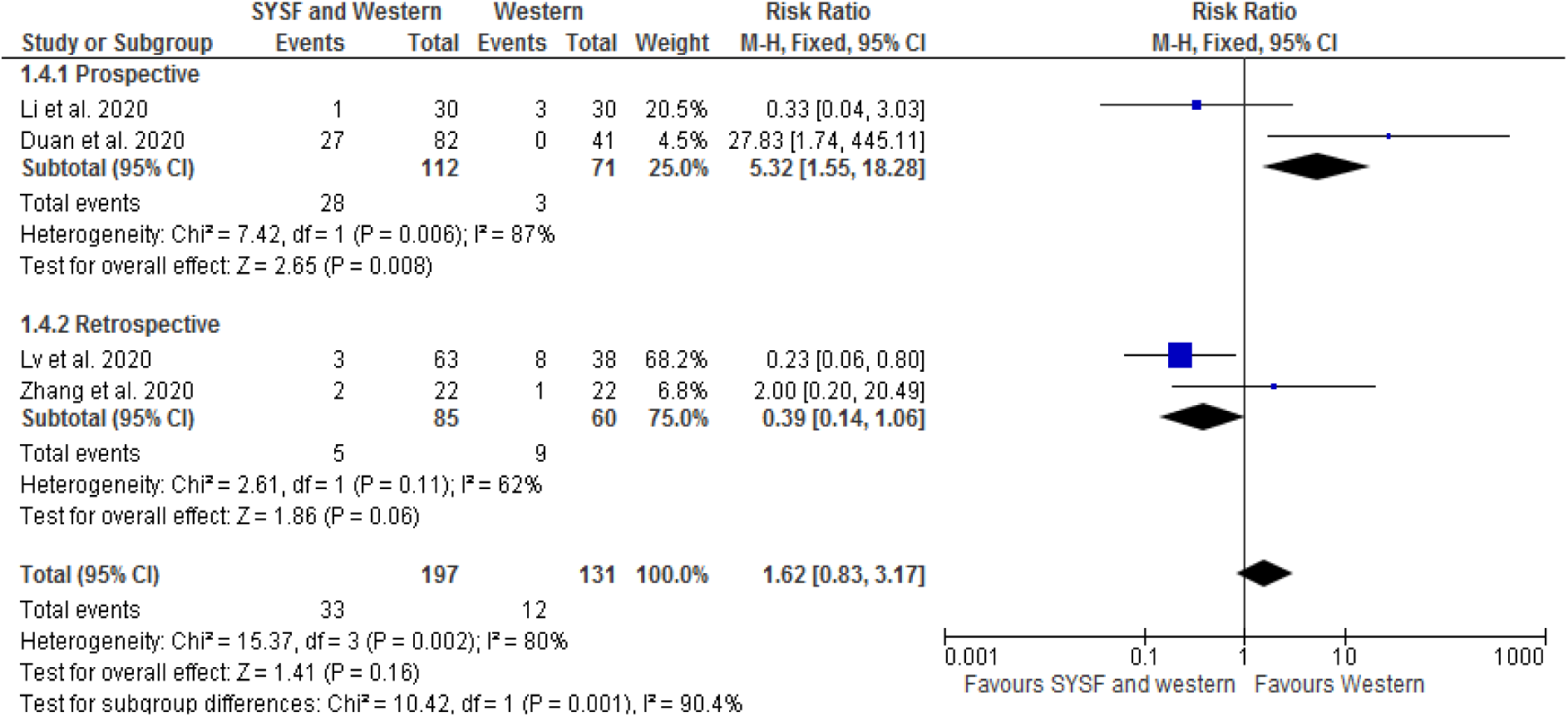
Adverse events during the management of patients

In addition, we performed subgroup analyses of prospective and retrospective studies separately.

The prospective studies analysed were Li et al., 2020 and Duan et al., 2020 comprising 183 patients, 112 in the SYSF and Western medicine arm, and 71 in the Western medicine alone arm. This gave a statistically significant risk ratio of 5.32 (95% CI 1.55 to 18.28, P = 0.006) in favour of the Western medicine alone arm, with an I2 of 87% (Fig. 7), suggesting that patients participating in studies performed prospectively who received SYSF and Western medicine reported more adverse events.

The retrospective studies analysed were Lv et al., 2020 and Zhang et al., 2020 comprising 145 patients, 85 in the SYSF and Western medicine arm, and 60 in the Western medicine alone arm. This gave a risk ratio of 0.39 (95% CI 0.14 to 1.06, P = 0.06) with an I2 of 62% (Fig. 7), suggesting that there is no statistically significant difference in adverse events reported by patients in the two arms in studies performed retrospectively.

##### 5. Duration of hospitalisation

Two studies reported this outcome and were analysed - Li et al., 2020 and Zhang et al., 2020.

The total population analysed comprised 96 patients, 49 in the SYSF and Western medicine arm, and 47 in the Western medicine alone arm. This gave a mean difference of -0.73 (95% CI -5.19 to 3.73, P = 0.75) with an I2 of 91% (Fig. 8), suggesting that there is no statistically significant difference in the duration of hospitalisation between the two arms.

**Figure 8.**
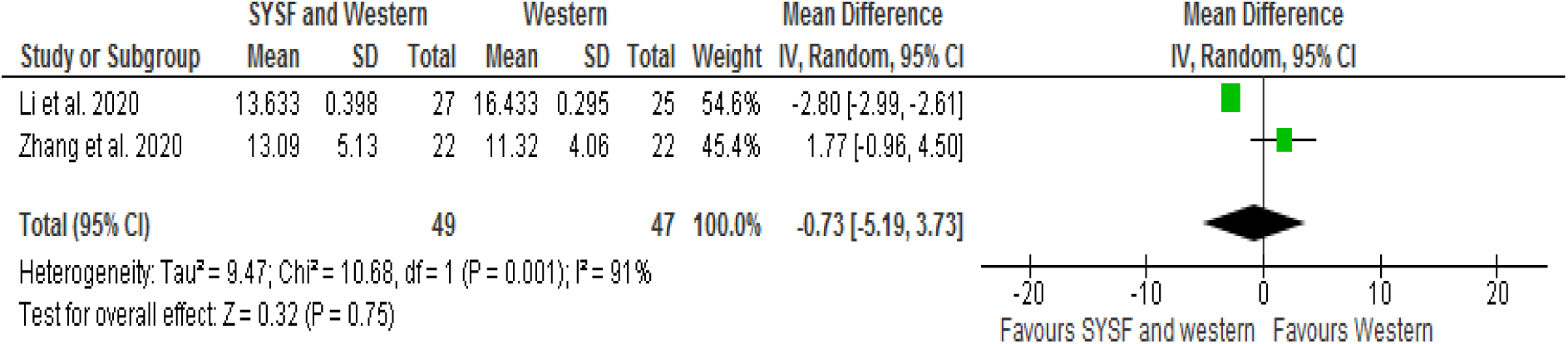
Hospitalization duration between combined therapy vs Western treatment alone

## Discussion

This review evaluates the effectiveness of SYSF in the management of COVID-19 symptoms. We describe the individual components of SYSF as shown in table 1.

### Formula J

In this review, it showed that Formula J combined with conventional treatment of COVID-19 can effectively improve fever, cough, fatigue, sputum production, and relieve patient anxiety. (Table 2). Looking at hospitalisation rate and improvement of other symptoms such as headache, sore throat, rhinorrhoea and nausea, there was no statistical difference between the treatment group and the control group. However, there is a significant difference in regards to adverse events experienced by the treatment group. 32.39% of the patients in the treatment group developed diarrhea symptoms in this study, as compared to 0% of patients in the control group experiencing adverse events. This may be due to the high dose of Formula J used in this study (2 sachets of 5g each time, 3 times a day), as compared to the usual dose of 1 sachet of 5g each time, 3 times a day.^32^ There is no research comparison of the degree of adverse reactions caused by high and low doses of Formula J, and whether diarrhea is an adverse reaction of Formula J is still to be further discussed. Although Formula J may meet current treatment needs, it should be pointed out that there are still deficiencies as patients in the study were not treated according to syndrome differentiation. Treatment using Formula J based on syndrome differentiation is a potential direction for further research.

### Formula L

Comparing 4 different papers on Formula L (Table 3), the treatment groups in all 4 papers demonstrated a faster recovery of fever, cough and fatigue. Formula L was also effective in the normalization of inflammatory markers like CRP and ESR, as well as both lymphocyte and neutrophil count. Importantly, Lv et al showed no statistical difference in terms of adverse events between the treatment and control groups. The adverse events that patients in the treatment group experienced are diarrhoea and loss of appetite which subsequently resolved with the stoppage of the management plan, with no long term sequelae. This is a promising first step into the safety aspect of Formula L, and future research can focus in this direction in order to establish the full safety profile of Formula L.

### Formula Q

Li et al showed that Formula Q with conventional antiviral management led to significant improvement in terms of resolution of symptoms such as cough and fever (Table 4), and decreased hospitalisation duration as compared to the control group. Resolution of CT Thorax findings was also quicker in the treatment group. There were no statistical differences in the worsening of symptoms between the treatment and control groups, even though the treatment group had fewer patients deteriortating (6 patients in treatment group; 12 patients in control group). Looking at the safety profile of Formula Q, one patient experienced nausea during treatment. This was resolved after the introduction of pantoprazole into the management of this patient.

### Formula Xb

Zhang et al demonstrated significant rates of recovery and resolution of CT thorax in the treatment group. (Table 5) There was however, no significant difference in resolution of inflammatory markers and nucleic acid conversion rate across the both groups after treatment. As the criteria for effective recovery stated in the study is defined as an improvement in symptoms, nucleic acid conversion and resolution of CT Thorax, it would suggest that the recovery rate in the treatment group is limited by the nucleic acid conversion rate. In addition, there was no significant difference in adverse events as compared to the control group. As such, it would be an interesting research direction to study the mechanism of action of Formula Xb, especially on its potential impact on ARDS in other diseases.

### Overall analysis of SYSF and Western treatment

The combination of Formula L and Western interventions led to a speedier and more favourable outcome in the treatment of COVID-19 patients, specifically for symptoms like fever, cough and fatigue. (Fig. 2-4) Low heterogeneity was observed in the three studies done.

In addition, the combination of Formula L or Q with Western interventions resulted in a shorter duration of fever, and improvement of various other symptoms such as musculoskeletal pain, nausea, vomiting, rhinorrhoea and chest tightness. Similar results were not shown in Formula J and Xb. (Fig. 5-6)

In terms of overall safety profile, more research would have to be done in terms of interaction between SYSF and Western medications, specifically Formula J. The prospective study done by Duan et al showed a statistically significant increase in adverse events in the treatment group as compared to the control group. (Table 2) This might suggest possible interactions between Formula J and western medications.

The study done by Lv et al. showed a lower rate of adverse effects when Formula L was used in conjunction with western medications as opposed to a higher rate when Formula Xb was used demonstrated by Zhang et al.. (Fig. 7) However, as a whole, there was no statistical significant difference in adverse events between the combined SYSF and Western interventions compared to Western interventions alone. (Fig. 7).

There was no significant difference to the duration of hospitalisation between combined SYSF and Western interventions as compared to Western interventions alone. (Fig. 8) However when the data was further analysed, Formula Q (T: -2.80 days) as opposed to Formula Xb (T: +1.70 days) resulted in shorter stay within the hospital. Both papers reported a high degree of heterogeneity when comparing the duration of hospitalisation between the treatment and control group.

This review is limited by the lack of randomised control trials and prospective studies for each formula in SYSF. This is expected due to the ongoing and sudden pandemic of COVID-19, making trials difficult to conduct due to the lack of manpower available. This is compounded by new formulas that have been specifically concocted for COVID-19. As such, a mixed method review was employed, analysing both prospective and retrospective studies with a comparator. Even though there is high heterogeneity among the analysed papers due to different formulas being studied, we can use these papers to understand the overall effects of SYSF on patients with COVID-19 from multiple angles.

In this review, Formula L and Q showed promising results on the management of symptoms of COVID-19 patients with possibly lower levels of adverse events when combined with Western medications. Formula J and Xb do not appear to be effective, and might result in an increased number of adverse events and duration of stay when used. Further studies including a larger cohort size should be included to further evaluate the effects and safety of SYSF.

## Data Availability

NA

## Notes

**Competing Interest Statement:** All authors have completed the Unified Competing Interest form and declare: no support from any organisation for the submitted work; no financial relationships with any organisations that might have an interest in the submitted work in the previous three years, no other relationships or activities that could appear to have influenced the submitted work.

### Competing Interest Statement

The authors have declared no competing interest.

